# Making use of an App (Tawakkalna) to track and reduce COVID transmission in KSA

**DOI:** 10.1101/2022.10.16.22281142

**Authors:** Donal Bisanzio, Richard Reithinger, Sami Almudarra, Reem F. Alsukait, Di Dong, Yi Zhang, Sameh El-Saharty, Hala Almossawi, Christopher H. Herbst, Ada Alqunaibet

## Abstract

Since March 2020, the Kingdom of Saudi Arabia (KSA) has launched several digital applications to support the intervention response to reduce the spread of SARS-CoV-2. At the beginning of 2021, the KSA Government introduced a mandatory immunity passport to regulate access to public venues. The passport was part of the strategy of resuming public activities before reaching high vaccination coverage. The passport was implemented as a new service in the Tawakkalna mobile phone application (App). The immunity passport allowed access to public locations only for the users who recovered from COVID-19 or those who were double vaccinated. Our study aimed to evaluate the effectiveness of the immunity passport, implemented through the Tawakkalna App, on SARS-CoV-2 spread. We built a spatial-explicit individual-based model to represent the whole KSA population (IBM-KSA) and its dynamic on a national scale. The IBM-KSA was parameterized using country demographic, remote sensing, and epidemiological data. The model included non-pharmaceutical interventions and vaccination coverage. A social network was created to represent contact heterogeneity and interaction among age groups of the population. The IBM-KSA also simulated the movement of people across the country based on a gravity model. We used the IBM-KSA to evaluate the effect of the immunity passport on the COVID-19 epidemic’s outcomes. The IBM-KSA results showed that implementing the immunity passport through the Tawakkalna App mitigated the SARS-CoV2 spread. In a scenario without the immunity passport, the KSA could have reported 1,515,468 (95% confidence interval [CI]: 965,725-1,986,966) cases, and 30,309 (95% CI: 19,314-39,739) deaths from March 2021 to November 2021. The comparison of IBM-KSA results with COVID-19 official reporting estimated that the passport effectively reduced the number of cases, hospitalizations, and deaths by 8.7 times, 13.5 times, and 11.9 times, respectively. These results showed that the introduction of the immunity passport through the Tawakkalna App was able to control the spread of the SARS-COV-2 until vaccination reached high coverage. By introducing the immunity passport, The KSA was able to allow to resume most of public activities safely.

## Background

The response to the COVID-19 pandemic has been characterized by the use of several information, communication, and technology application s (ICT apps). Leveraging the widespread population coverage of smart phones and access to the internet, the pandemic has stimulated the development and launch of many digital e-health applications to support governments by mitigating COVID-19 outcomes [1]. In March 2020, Singapore; the Republic of Korea (Korea); Hong Kong SAR, China; and Japan deployed contact tracing ICT apps to support the contact tracing of SARS-CoV-2 cases [2]. Following the success of reducing the spread of the disease in countries where the contact tracing apps were employed, many other countries decided to deploy these apps to support interventions aiming to reduce the SARS-CoV-2 spread. These new e-health solutions used innovative technologies based on Bluetooth, Global Positioning System (GPS), artificial intelligence, and machine learning [3].

The aim of these ICT solutions was to improve health care system delivery and reduce virus circulation. Apps were created to provide information about COVID-19 both nationally and internationally, help to track virus exposure and confirmed cases, improve access to health services using remote-health solutions, regulate movement during lockdown and curfew periods, support vaccination campaigns, and regulate international travel and access to public spaces [3]. Most of the launched ICT apps were able to perform contact tracing, carry out self-assessments, book testing appointments, report on testing results, and allow for remote access to health services [4].

When COVID-19 vaccinations become available early 2021, countries decided to develop and launch ICT solutions to support the rollout of available vaccines that included an *immunity passport* (also called a *risk-free certificate*) feature. Apps—often linked to a country’s ministry of health or health providers’ data platforms—were created to book vaccination appointments and record the immunization status of people. Many countries adopted immunity passport systems to identify vaccinated people, allowing them to move more freely, both nationally and internationally, than non-vaccinated individuals [5]; adoption of these immunity passport apps was also used to increase vaccination coverage among the population. The immunity passport is not a new concept: this approach has been used for decades to reduce the risk of yellow fever infection in many countries with high transmission risk [6]. However, the effectiveness of these solutions has not been fully assessed, given the difficulty of untangling the role of each digital application on SARS-CoV-2 circulation and COVID-19 outcomes.

After the first case of COVID-19 was recorded in Saudi Arabia on March 3, 2020, the government rapidly adopted NPIs to mitigate the spread of the virus among its population. The rolled out NPIs included closing national borders, lockdowns, curfews, school closings, and enforcing mask-wearing and physical distancing in public spaces. By imposing strict NPIs, KSA recorded fewer than approximately 500 daily reported cases from September 2020 to the end of March 2021 [7]. Indeed, a recent study estimated that between June 2020 and June 2021, the adoption of these NPIs averted roughly 4 million cases, 600,000 hospitalizations, and 70,000 deaths due to COVID-19 [8].

Like other countries, KSA has implemented multiple ICT solutions to support the national health system in the response to COVID-19—a step that also built on and was aligned with KSA’s health system modernization set forth in the country’s Vision 2030 National Transformation Program [9]. The aim of the program was to increase access to health care by improving remote services (screening, prescription, scheduling visit appointments) for the population [9]. KSA was not new to using ICTs to handle emerging infection diseases. The country had already adopted ICT for handling the spread of Middle East respiratory syndrome coronavirus virus (MERS-CoV) in 2015 [10-11]. Increasing the e-health capacity of the health system resulted in an improved access to services with a high cost-benefit for the KSA health system [12]. Furthermore, as of January 2020, approximately 89 percent of the KSA population had access to the internet and 96 percent of the adult population owned smart phones [13]. Given the country’s experience with ICT to respond to health emergencies, its new public health strategy—based on remote solutions and the high penetration of information technology—helped KSA to rapidly deploy e-health solutions by adding new features to already-launched apps and developing new ones. Although many applications were launched from government and private sector [14], the pillars of the KSA e-health response to COVID-19 were five mobile phone applications: Mawid, Tabaud, Sehha, Tetamman, and Tawakkalna (Tables S1 and Table S2). Among all these applications, only the use of the Tawakkalna App was made mandatory for the KSA population.

The Tawakkalna app was launched in the KSA in April 2020. This app was designed to regulate people’s movements during lockdowns and curfews. The app records the health status of users and allows them to request movement permits during curfews and lockdowns. A color system also shows whether the user is able to travel freely in the country: green means that the user can travel; yellow means that the person has been exposed, could become positive, and needs to quarantine; and red means that the person is infected and has to remain in self-isolation. The number of app users rapidly increased during the month following its release, reaching about 20 million users by March 2021. The functions of Tawakkalna app were updated when vaccination was rolled out in December 2020 [15]. In January 2021, the MOH launched the Immunity Passport service as a new functionality of the app [16], which recorded the user’s COVID-19 vaccine doses received, the vaccine brand they received, and the date of vaccination. People who received two COVID-19 vaccine doses were then provided with access to public spaces and events. Furthermore, the app allowed KSA to fully resume national travel, reopen public venues, and resume public events starting in March 2021; it also allows for the resumption of in-person teaching for children above 11 years of age in August 2021 [17].

The use of the Tawakkalna app as an immunity passport, in conjunction with mandatory enforcement of NPIs, has been the country’s main strategy for re-opening shops, restaurants, and other public venues, and for allowing public events. The use of the app also aimed to mitigate the spread of the virus while, at the same time, reopening public activities before the country reached a high vaccination coverage rate. Furthermore, the app was the only ICT made mandatory for the KSA population.

Estimating the impact of using the Tawakkalna app as an immunity passport is crucial to better understanding the role of implementing such a policy to minimize future outbreaks of COVID-19 or other highly transmissible infections. The aim of this study was to use a novel epidemiological individual based modeling framework (IBM-KSA) representing KSA’s population to assess the effect of Tawakkalna app as an immunity passport, using the estimated number of cases, hospitalizations, and deaths from March 2021 to November 2021 as its metric. The epidemiological model had already been used to estimate the health benefit of NPIs adopted by KSA before the introduction of the vaccines [8].

## Method

### IBM-KSA model structure description

The impact of using the Tawakkalna app as a COVID-19 immunity passport for tracking SARS-CoV-2 circulation in the Kingdom of Saudi Arabia (KSA) was modeled by simulating interaction at the person level using an individual-based model. The individual-based model used to represent the KSA population was named IBM-KSA. The model had already been used to estimate the effect of non-pharmaceutical interventions (NPIs) adopted during the first phase of the pandemic, between March 2, 2020, and July 31, 2021 [8]. The IBM-KSA model utilizes a contact network structure to simulate interactions among the entire KSA population—that is, among approximately 34 million people. The network structure was preferred to an agent model because it allows for contact heterogeneity among people while reducing computational load. The model was built using demographic data to reproduce a population with characteristics of the actual KSA population. The IBM-KSA also accounts for differences in contact interactions among age groups [18-21] as well as accounts for movement flux among cities—a main driver of SARS-CoV-2 spread in a country [22].

### COVID-19 Measures between March and November 2021

The model included NPIs and the vaccination campaign, as their implementation dates as of November 2021. The model also included the introduction of the immunity passport through the Tawakkalna app to calibrate the model with real data. After calibration, the IBM-KSA simulated projections of COVID-19 cases, hospitalizations, and deaths, with or without the introduction of the immunity passport. The modeled scenarios simulated the epidemic from March 1, 2021, to November 30, 2021. This time window was chosen to capture the effect of the use of Tawakkalna app immunity passport before the introduction of the SARS-CoV-2 Omicron variant in the country.

### Epidemiological Model

The transmission dynamic of COVID-19 was represented in the model by the classical transition status seen in SEIR compartmental models:

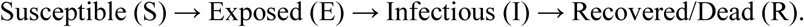

COVID-19 national data were used to estimate the hospitalization and fatality rates for KSA [7] to determine the number of infected individuals who would also be at risk of being hospitalized or dying. The transition from one status to another is a function of pathogen characteristics (for example, virulence, incubation period, infectious period, hospitalization, and fatality rate); interaction among individuals (only for S through I); and immunization status [22-27]. The IBM-KSA model also accounts for the fraction of asymptomatic cases, set as equal to 80 percent [27]. A detailed description of the IBM-KSA model framework and transmission parameters can be found in Bisanzio et al. [13]. Here follows a list of new and updated parameters used to modify the KSA-IBM published in Bisanzio et al. [13].

Transmission parameters:

▪ Transmission probability per hour: 0.06 and 0.12 for Alpha and Delta variants, respectively [8, 28]
▪ First case of Delta variant reported in August 2021 [29]
▪ Naturally acquired immunity: >12months (assumption) but with reduced protection over time [30]
▪ Hospitalization rate equal to 10 percent and death rate equal to 2 percent of infected individuals [7]

NPIs included in the IBM-KSA:

▪ Mask wearing for 70 percent of the population and social distancing for 80 percent of the population Full isolation for 80 percent of the population [8]
▪ Resumed in-person education only for students 11 years of age with the following restrictions:
  ▪ Mask wearing and social distancing set to 90 percent
  ▪ Class isolation in case of detected cases among students
  ▪ Children can attend class only if double-vaccinated

Vaccination status of KSA population:

▪ 70 percent of the population was fully vaccinated by November 2021
▪ 90 percent of 11–17 age group was fully vaccinated by November 2021

Vaccination campaign:

▪ ∼350,000 vaccinations per day [31]
▪ 3-week lag between the two doses
▪ Vaccination offered to people > 17 years old
▪ Vaccination for people 11–17 years of age began June 2021
▪ Maximum coverage of the population set to 95 percent
▪ Delay of second dose for different age groups from April to July 2021

## Results

The KSA-IBM results showed that using the Tawakkalna app as an immunity passport was able to substantially mitigate the spread of COVID-19 in the country. If the immunity passport had not been in place, the number of cases would have drastically increased after the easing of NPIs at the beginning of March 2021. Additionally, the number of cases would have peaked in June 2021, with more than 600,000 (95% CI: ∼400,000–∼900,000) cases per week (Figure 1). The estimated number of cases fell after June as a result of the expanded rollout of vaccination that also targeted children ages 11–17. The number of reported cases showed a slight increase until February 2021, but then it remained stable, with roughly 8,400 cases per week from the beginning of May 2021 to the end of August 2021 (Figure 1). The massive reduction of cases after August could be linked to the high proportion of population having received their second vaccine. The number of cases stabilized until the end of November 2021, with approximately 300 cases per week (Figure 1).

**Figure 1.**
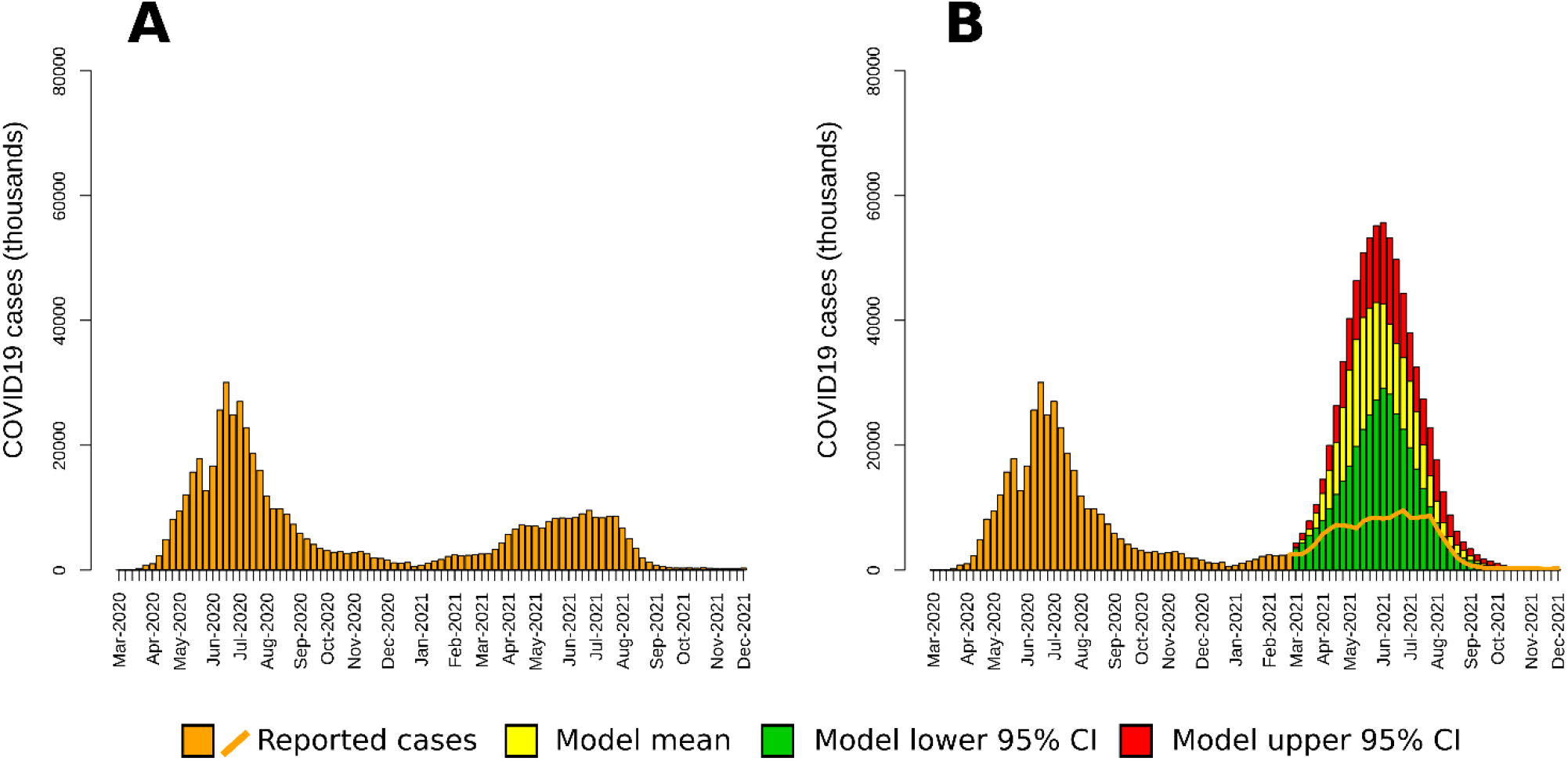
Epidemic Curves of the COVID-19 Epidemic in KSA with an Immune Passport (panel A) and Scenarios without an Immune Passport (panel B), March 1, 2020, to December 1, 2021. The yellow bars are the mean values of the simulation, and the green and red bars are the 95 percent confidence interval (95% CI). The orange bars and line represent the reported number of cases until December 1, 2021 [12]. CI = confidence interval.

The results of the model showed that the app was able to reduce the spread of the virus while vaccination coverage was increasing, reaching a threshold after which the spread of the virus was reduced. Using the IBM-KSA model, we estimate that the use of the app was able to avert 1,343,099 cases, 112,322 hospitalizations, and 27,967 deaths from March to November 2021 (Table 1). The comparison of IBM-KSA results with COVID-19 official reporting estimated that the passport effectively reduced the number of cases, hospitalizations, and deaths by 8.7 times, 13.5 times, and 11.9 times, respectively (Table1). After August 2021, when most of the population was fully vaccinated against SARS-CoV-2, the app’s effectiveness was reduced because an important fraction of the population was—due to their vaccination status—no longer affected by the limitations imposed by the app (Tables 1 and 2). We estimated that dropping the use of Tawakkalna after reaching high fraction of fully vaccinated people in the population could have increase the number of cases, hospitalizations, and deaths by by 1.8 times, 1.7 times, and 1.3 times, respectively (Table2).

**Table 1.**
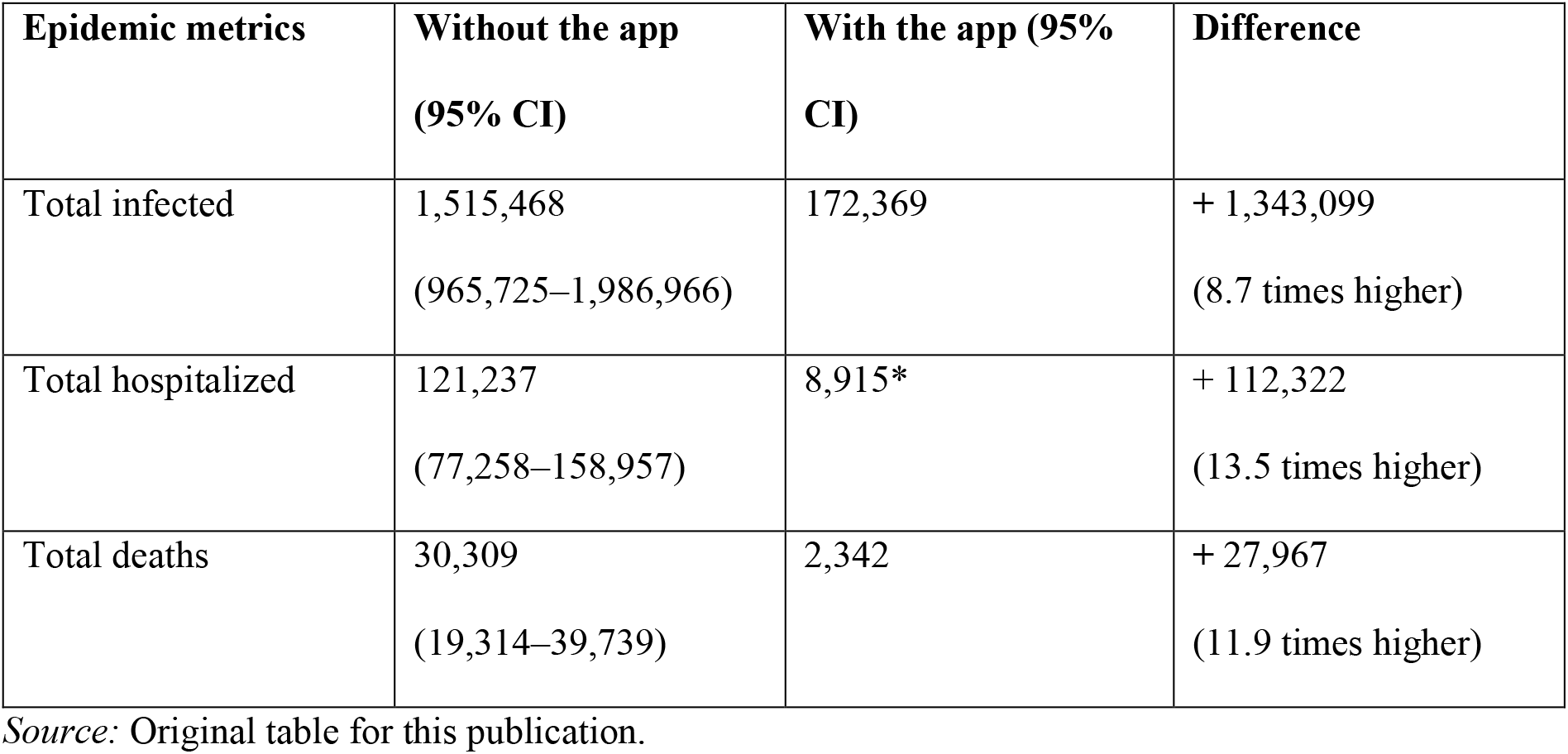
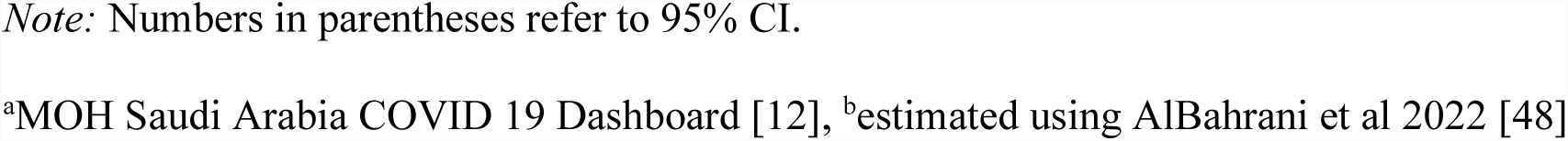
The Impact of Using the Tawakkalna App as an Immunity Passport on Cases, Hospitalizations, and Deaths, March 2021 to November 2021.

| Epidemic metrics | Without the app<br>(95% CI) | With the app (95%<br>CI) | Difference |
| --- | --- | --- | --- |
| Total infected | 1,515,468<br>(965,725–1,986,966) | 172,369 | + 1,343,099<br>(8.7 times higher) |
| Total hospitalized | 121,237<br>(77,258–158,957) | 8,915* | + 112,322<br>(13.5 times higher) |
| Total deaths | 30,309<br>(19,314–39,739) | 2,342 | + 27,967<br>(11.9 times higher) |

Source: Original table for this publication.
*Note:* Numbers in parentheses refer to 95% CI.
<sup>a</sup>MOH Saudi Arabia COVID 19 Dashboard [12], <sup>b</sup>estimated using AlBahrani et al 2022 [48]

**Table 2.** The Impact of Using the Tawakkalna App as an Immunity Passport on Cases, Hospitalizations, and Deaths, August 2021 to November 2021.

| Epidemic metrics | Without the app<br>(95% CI) | With the app (95%<br>CI) | Difference |
| --- | --- | --- | --- |
| Total infected | 38,847<br>(26,571–62,996) | 23,426<sup>a</sup> | + 15,421<br>(1.7 times higher) |
| Total hospitalized | 3,107<br>(2,125–5,039) | 1,639<sup>b</sup> | +1,468<br>(1.8 times higher) |
| Total deaths | 776<br>(531–1,259) | 593<sup>a</sup> | + 183<br>(1.3 times higher) |

<sup>a</sup>MOH Saudi Arabia COVID 19 Dashboard [12], <sup>b</sup>estimated using AlBahrani et al 2022 [48]

## Discussion

The IBM-KSA model’s results showed that introducing the Tawakkalna app immunity passport was able to reduce the circulation of SARS-CoV-2, resulting in a low number of reported cases, hospitalizations, and deaths. Re-opening without the passport would have resulted in over a million more reported cases, over 100,000 more hospitalizations, and around 30,000 more deaths from March 2021 to November 2021 than with the passport. Furthermore, the introduction of the immunity passport allowed the country to maintain lower circulation of the virus while vaccination coverage was increasing. These findings are consistent with studies performed in other countries introducing a national immunity passport. The introduction of the immunity passport (called the *green pass*) in Italy was linked to a reduction of reported COVID-19 cases, hospitalizations, and deaths, and reduced the virus’s transmission in the early stage of the vaccination campaign [32].

Making the app mandatory probably improved the impact of the app on COVID-19 transmission. Studies evaluating ICT solutions that allow for contact tracing have found that when use is non-mandatory, their effect on disease transmission is limited. First studies investigating contact tracing apps have shown that such apps successfully reduced the incidence of COVID-19 in England, Switzerland, and Hong Kong SAR, China [33-35], with the apps’ success linked to the percentage of the population using them. In contrast, in Australia, a study showed that a contact tracing system through an app had a small and lower-than-expected effect on COVID-19 incidence [36], which was likely the result of the low number of app users (only approximately 22 percent of the population) [36]. These results are consistent with a recent systematic review showing only weak evidence of the impact of contact tracing on COVID-19 cases [37]. A systematic review highlighted the way modeling studies had estimated high effectiveness of contact tracing apps only with a high level of population uptake (higher than 50 percent) [38]. The non-mandatory policy of contact tracing apps could be at the base of low population uptake in many countries, resulting in a dimmed efficacy of this tool.

The use of an immunity passport that reduces the movement of the non-immunized fraction of the population could also result in a rapid increase of vaccination coverage. Enforcing the need to prove their immunization status to access workplaces, shops, schools, public venues, and events would make people keener to be vaccinated. A recent study showed that introducing an immunity passport could speed up the vaccination campaign and provide high vaccination coverage in less time than countries without a mandatory passport [39]. Thus, the adoption of an immunity passport could benefit vaccination campaigns by minimizing the circulation of the virus during the period when vaccination coverage is still increasing, and boost vaccination adherence.

Although the introduction of an immunity passport has been criticized for its ethical implications [40], as of October 2021, 144 countries have implemented different forms of immunity passports reporting people’s immunization status to regulate access to public space and people movement or to international travel [5]. Depending upon country regulations, the passport could show vaccination status, proof of a negative COVID-19 test, proof of recovery from the virus, or a combination of such information.

Because updating digital immunity passport apps through mobile phones is so easy, immunity passports have also been used to reduce the spread of the new SARAS-CoV-2 Omicron variant, first reported in South Africa in November 2021 [41]. After Omicron was detected in KSA, the number of reported COVID-19 cases rapidly increased and the country re-established mandatory mask-wearing and social distancing. The country’s Ministry of Health (MOH) also decided to update the rules to allow access to public spaces only for those people who have received a third dose of the vaccine (the booster dose) [42]. These rules were able to swiftly decrease the number of cases, avoiding major disruption to the health system and mitigating the outcome of Omicron spread [7].

In conclusion, the study showed that introducing an immunity passport through the Tawakkalna app could result in high public health benefits by reducing the burden of COVID-19. Such immunity passports are a tool that can easily be modified to face new public health emergency threats—such as the emergence of new SARS-CoV-2 variants or future pandemics—and to implement changes in the public health response. Adopting a digital immunity passport, such as the Tawakkalna app, can reduce the need for curfews and lockdowns; the result is a positive health impact that also protects the country’s economy by allowing people to safely resume economic activity and international traveling and protecting human capital.

## Data Availability

All data produced in the present study are available upon reasonable request to the authors

## Aknowledges

The study was developed by the Saudi Public Health Authority, with technical support from individuals from RTI International, and the World Bank. The authors are grateful for the support provided by Rekha Menon (Practice Manager, Health Nutrition and Population, Middle East and North Africa region, World Bank) and Issam Abousleiman (Country Director, GCC countries, World Bank). The findings, interpretations, and conclusions expressed in this work are those of the authors and do not necessarily reflect the views of the authors’ employers or organizations that funded the work.

