## Supplementary material for "Making use of an App (Tawakkalna) to track and reduce COVID transmission in KSA": Tables S1 and Table S2

**Table 1. Overview of the E-Health Mobile Phone Apps Existing before COVID-19**

| **App name** | **Pre COVID-19** | **COVID-19 adaptation** | **Logo** |
|---|---|---|---|
| *Sehhaty* | Following the 2018 KSA strategy to increase access to health care services, an e-consultation app based on artificial intelligence (AI) algorithms, called *Sehhaty*, was deployed alongside Mawid (Alharbi, Alzuwaed, and Qasem 2021). While the Mawid app allows the user to schedule physical appointments with health facility service providers, the Sahhaty app provides easy access to remote medical consultations via video calls with doctors through smartphones (Aldekhyyel, Almulhem, and Binkheder 2021). The Sehhaty app also applies AI to improve users' experience by providing targeted health information based on survey assessments. | The KSA health system has been under pressure during the pandemic not only because of the high load of cases needing hospitalization, but also because of the high proportion of health workers who were exposed to infected subjects. The Sahhaty app allows pressure on health facilities to be reduced and also reduces health workers' exposure to the virus. | |
| *Mawid* | The Mawid app was developed to improve access to appointments by allowing remote access to the population. The app was launched in May 2019 as one of the ICT tools in the health system's restructuring and reforms. Using this app, users are able to access 98 percent of hospitals and primary care centers across the country directly from their phones. | When the COVID-19 epidemic began in KSA, the national health system had already a mobile-phone app called *Mawid* to manage the central diagnostic testing appointment system. At the beginning of the COVID-19 KSA epidemic, Mawid's self-assessment tool was updated, allowing suspected COVID-19 cases to schedule appointments at COVID-19 testing facilities (Alanzi et al. 2022). The app was also modified to provide information about COVID-19 awareness. | |

*Source:* Original table for this publication.

**Table 2. Overview of the E-Health Mobile Phone Apps Developed after COVID-19**

| | |
|---|---|
| *Tabaud* | As did many other countries (Hidayat-ur-Rehman et al. 2021), KSA deployed a contact tracing app—called *Tabau*—to control the spread of SARS-CoV-2. This application uses Bluetooth technology to notify the user when they come in close proximity with other users who have reported being infected. For all possible exposed users, the app calculates the risk of infection given phone proximities with an infected user. All infection data uploaded through the app are double-checked by a connection with the Ministry of Health (MOH) COVID-19 data set system. By recording users' mobility and proximity to one another, the Tabaud app is able to inform users if they were in close contact with infected users during the previous 14 days. The app also provides information to all exposed users on preventive measures to take (for example, self-isolation, symptom checking) in order to minimize virus spread. |

| | |
|---|---|
| *Tetamman* | The Tetamman app was launched in April 2020 with the aim of supporting individuals who are in self-isolation or quarantine. The app can book COVID-19 tests, access COVID-19 testing results, and report daily on symptom status. The app is also able to book an appointment for an examination and request necessary medical assistance (Hidayat-ur-Rehman et al. 2021). |
| *Tawakkalna* | In April 2020, KSA launched Tawakkalna, an app designed to regulate people's movements during lockdowns and curfews. The app records the health status of users and allows them to request movement permits during curfews and lockdowns. A color system also shows whether the user is able to travel freely in the country: green means that the user can travel; yellow means that the person has been exposed, could become positive, and needs to quarantine; and red means that the person is infected and has to remain in self-isolation. The number of app users rapidly increased during the month following its release, reaching about 20 million users by March 2021. |
